# Household bubbles and COVID-19 transmission: insights from percolation theory

**DOI:** 10.1101/2020.12.16.20248311

**Authors:** Leon Danon, Lucas Lacasa, Ellen Brooks-Pollock

**Affiliations:** University of Exeter College of Engineering Mathematics and Physical Sciences, CEMPS North Park Road, Exeter, EX4 4QF, United Kingdom; Queen Mary University of London, School of Mathematical Sciences, Mile End Road London, E1 4NS, United Kingdom; University of Bristol School of Veterinary Sciences Langford, BS40 5DU, United Kingdom; University of Bristol, Population Health Sciences, Bristol Medical School, BS8 2BN, United Kingdom

## Abstract

**Background:** In the era of social distancing to curb the spread of COVID-19, bubbling is the combining of two or more households to create an exclusive larger group. The impact of bubbling on COVID-19 transmission is challenging to quantify because of the complex social structures involved.

**Methods:** We developed a network description of households in the UK, using the configuration model to link households. We explored the impact of bubbling scenarios by joining together households of various sizes. For each bubbling scenario, we calculated the percolation threshold, that is, the number of connections per individual required for a giant component to form, numerically and theoretically. We related the percolation threshold to the household reproduction number.

**Results:** We find that bubbling scenarios in which single-person households join with another household has a minimal impact on network connectivity and transmission potential. Ubiquitous scenarios where all households form a bubble are likely to lead to extensive transmission that is hard to control. The impact of plausible scenarios, with variable uptake and heterogeneous bubble sizes, can be mitigated with reduced numbers of contacts outside the household.

**Conclusions:** Bubbling of households comes at an increased risk of transmission, however under certain circumstances risks can be modest and could be balanced by other changes in behaviours.

## Introduction

Transmission of close contact infections, such as COVID-19, fundamentally depends on social interactions between individuals. Patterns of social contact determine the rate and extent of spread in a population. Social networks are complex and hierarchical due to how society is structured. Social distancing has been one of the main methods for controlling SARS-CoV-2 transmission in the absence of a vaccine or effective pharmaceutical interventions. In the UK, social distancing measures have sought to limit social interactions outside households, due to the inherent challenges of preventing household transmission. Limiting all non-household contacts should reduce the reproduction number to close to zero, however in practice not all external contacts can be stopped.

Social distancing comes at a cost which disproportionately affects some individuals more than others. Detrimental effects include increased loneliness, social isolation, lack of physical and emotional support and reduced childcare provision, which can all be associated with a financial cost. The formation of ‘bubbles’, defined as small, non-overlapping, groups of households that are permitted to come into contact with each other, are intended to maintain benefits of social distancing while reducing the negative impacts of isolation. This, in essence, has the effect of creating one large household out of two or more smaller households. Social support bubbles have been used in various forms in countries including New Zealand, Belgium and the UK.

The precise definition of an allowed bubble has varied over time and between countries, making their effectiveness as a strategy difficult to quantify. The epidemiological impact of bubbles depends on rates of transmission and mortality, infrastructures for tracking, testing and isolating cases, protective work environments and safe transport modes for key workers. In the UK, support bubbles are bubbles of two households where one household contains a single adult or young children.

Childcare bubbles are bubbles where a person from one household provides childcare for another household. Christmas bubbles are groups of three households who are permitted to mix closely between 23 December and 27 December 2020. In all cases, the bubbles must be exclusive and non-overlapping and contacts within a bubble should be treated as household contacts if someone in the bubble tests positive for COVID-19 (1).

In this work we explore the impact that bubbling strategies have on the ongoing COVID-19 epidemic, to provide an evidence base and inform decisions. We use the distribution of household sizes in the UK in order to identify the UK conditions that will enable ‘bubbles’ to be safe and effective

## Methods

### Percolation theory

Percolation theory has served as a direct analogy for infectious disease transmission. Originating in material science to describe the flow of liquid through a porous medium, percolation theory has developed through mathematical abstraction to map epidemic transmission models on a social network to the purely structural problem of percolation of an underlying network. The epidemic threshold, above which an epidemic is very likely, is deeply related to the concept of the percolation threshold, above which a percolating cluster appears with a high probability (2).

Whereas mathematical models have usually focused on network abstractions (lattices or random graphs), in the last 20 years compelling empirical evidence has made it clear that the connection topology of real systems, such as household networks, are very different from the idealised lattices or random graphs traditionally studied in percolation theory. Instead, realistic networks display stylised patterns midway between order and disorder (3). and understanding how not only epidemics spread (4) but also how percolation can take place in models of complex networks with realistic characteristics has been the subject of intense study (see (5,6) and references therein).

### Constructing networks of households

To build the contact network of households, initially we consider the realistic household size distribution from the UK 2011 census (see Figure 1a) (7). Then, we assume that all individuals in a household form a clique (a fully connected subgraph) and consider each household as a single node in the network, effectively going from a network of individuals to a network of households. To generate between-household connections, we assume that each individual in a household has the same probability of generating external connections, therefore that the degree of a household-node is proportional to its size (number of individuals). We therefore say that a household with *n* individuals is of size *n* and thus constitutes a node with degree *n*. As a baseline, we finally assume that individuals are connected to individuals in other households at random, and accordingly build connections between household-nodes. This process of building the network of households is akin to a configuration model where the degree sequence of household-nodes is fixed (8) (Figure 1b). This is termed the *baseline network* without household bubbles.

**Figure 1:**
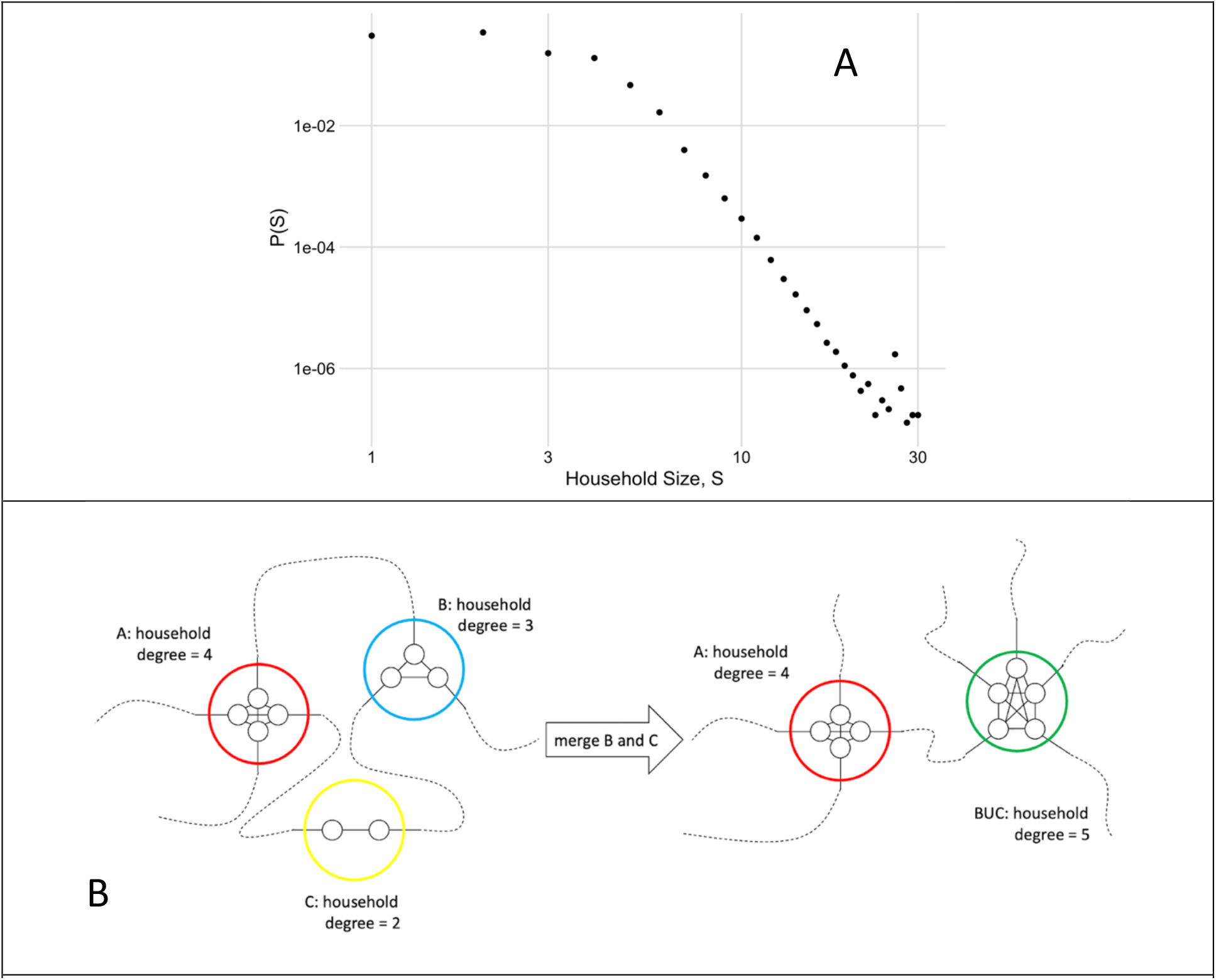
Schematic of network construction, bubbling and percolation analysis. A) The distribution of household sizes from the Office of National Statistics Census in 2011. B) A schematic of the formation of a network at random and merging of households

### Simulating the effect of bubbling

We modelled bubbling by combining multiple households into larger ones, thus increasing the number of external links. Once the bubbled network has been formed, we study how resilient it is against random removal of these external links (see next section on percolation analysis), that will in turn allow us to associate a certain level of transmission risk to a specific bubbling scenario. We considered scenarios where all households behave in the same way, as well as more plausible scenarios with variable take-up and behaviours:

- **2-bubbles**: All households join together with one other household, chosen at random, to create a bubble of two households.
- **3-bubbles**: All households join together with two other households, chosen at random, to create a bubble of three households.
- **1+1**: All single-person households (size 1 households) join together with one other single person household to make a two-person household.
- **1+n**: All single person households join together with another randomly chosen household of any size.
- **2+n**: All households of size 1 or 2 join with another household of any size.
- **Plausible best case:** 33% of households form a bubble; half are 2-bubbles and half are 3-bubbles.
- **Plausible reasonable case:** 50% of households form a bubble; half are 2-bubbles and half are 3-bubbles.
- **Plausible worst case:** 75% of households form a bubble; half are 2-bubbles and half are 3-bubbles.

### Percolation analysis

A component of a network is defined as a connected subgraph, i.e. as a set of nodes where each node is reachable to and from any other node following existing links. When the largest connected component of a network contains a finite fraction of the nodes of the network, we say the network has a giant component. Of course, all finite networks have a giant component (simply, the largest), but the concept reaches conceptual relevance in the limit of large networks.

In order to measure the robustness of networks before and after bubbling, we now perform a percolation analysis on the resulting networks, which essentially consists of measuring the proportion of links that need to be removed to break up the giant component. To do this, we begin by removing a proportion, (1 − *p*), of links from the network at random for all values of *p* from 1 to 0, decreasing in increments of 0.01. As (1 − *p*) is progressively increased, more between-household links are removed from the baseline network and the giant component decreases in size. At a critical point, *p*_*c*_(the so-called percolation threshold) the network fragments abruptly and the giant component disappears. We use *p*_*c*_ as the indicative variable: a network with a low percolation threshold is much more risky with respect to a disease propagating over it, as only a handful of additional links are needed to get from the non-percolating to the percolating phase.

For *p* ranging in [0,1], we also measure the number of households in the giant component and the *average size* of the other components in the network (also known as the order parameter). As soon as the giant component emerges, it quickly accrues most of the nodes of the network, meaning that the rest of components will be smaller. A discontinuity (peak) in the profile of this order parameter indicates the location of the phase transition, i.e. the threshold that distinguishes the non-percolating and percolating phases^1^. We repeated this procedure for each network following different bubbling scenarios, estimating the location of the threshold for each case. Since each realization of the network is stochastic, we repeated this 100 times for each scenario, and assessed the effects of intrinsic noise in the system.

As described above, the relative location of this phase transition is used to infer the relative impact of specific bubbling strategies. For a lower percolation threshold, the underlying network is more resilient to link removal; in other words, for the same per link probability of infection, transmission on a network with a low percolation threshold will affect a larger proportion of the population than a network with a high percolation threshold (see figure 2). Moreover, if the system is at the critical point on the *baseline network*, the relative position of the threshold on equivalent networks under different bubbling assumptions will indicate whether a strategy is better (subcritical, no giant component) or worse (supercritical, large giant component).

**Figure 2:**
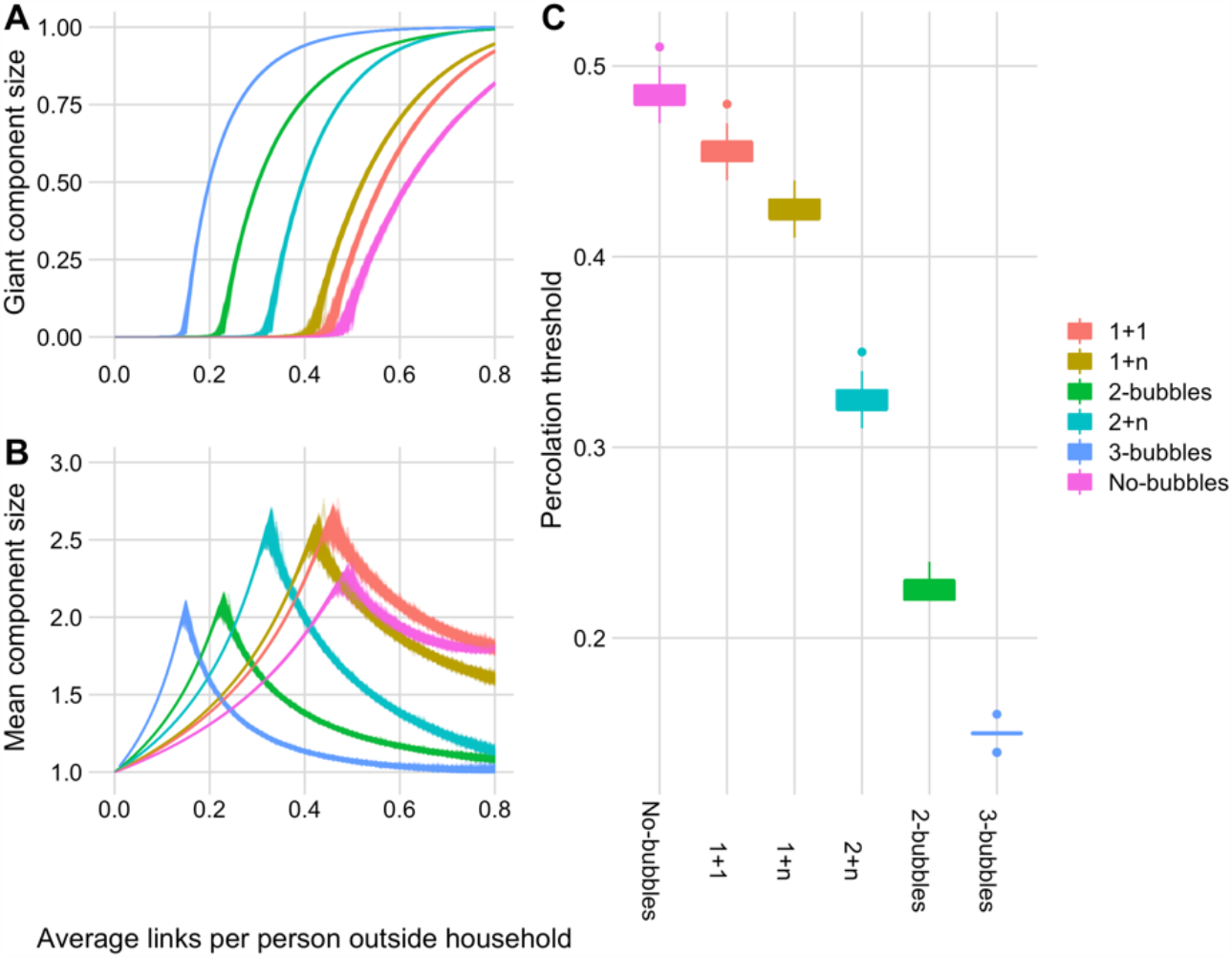
Percolation analysis for hypothetical bubbling scenarios. A) The proportion of households connected to the giant component. B) The mean component size (order parameter) for the same bubbling strategies. C) The percolation threshold for the different bubbling assumptions.

### Relating percolation thresholds to transmission potential

The percolation threshold is the point at which the giant component emerges (or disappears), and the network is globally connected (or disconnected). For locally tree-like networks, pc can be expressed in terms of the degree and degree fluctuations of the network:

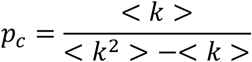

This purely topological property is in turn related to transmission potential and the reproduction number in the population(4,9).

Much of the focus in describing COVID-19 transmission has centred on the reproduction number, or R, defined as the average number of secondary cases caused by an average infected individual. However alternative reproduction numbers can be used to understand transmission at different scales, for example the household reproduction number, which is the average number of secondary households infected by an average household (10,11).

The household reproduction number is given as

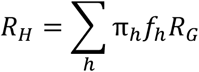

where *f*_*h*_ is the expected number of infections within a household of size *h, π*_*h*_ is the proportion of households of size *h* and *R*_*G*_ is the mean number of out-of-household infections by a single infected person (10). In our formulation, *R*_*G*_ = *p*

and *f*_*h*_ is *h* multiplied by the Secondary Attack Rate within households. We use the estimated Secondary Attack Rate from UK data of 40%-50% (12). Furthermore, below the percolation threshold, *R*_*H*_ = 0 because the infection is necessarily limited to small disconnected clusters.

## Results

With no household bubbling, we estimate a percolation threshold of 0.48, meaning that on average, if one out of every two people has a social contact outside of their household then a giant component emerges in the network of households and there is the potential for large-scale outbreaks. At the percolation threshold, the household reproduction number is estimated to be 0.47 - 0.59 (Figure 2), therefore even though a giant component exists transmission is not self-sustaining (Figure 5).

**Figure 3:**
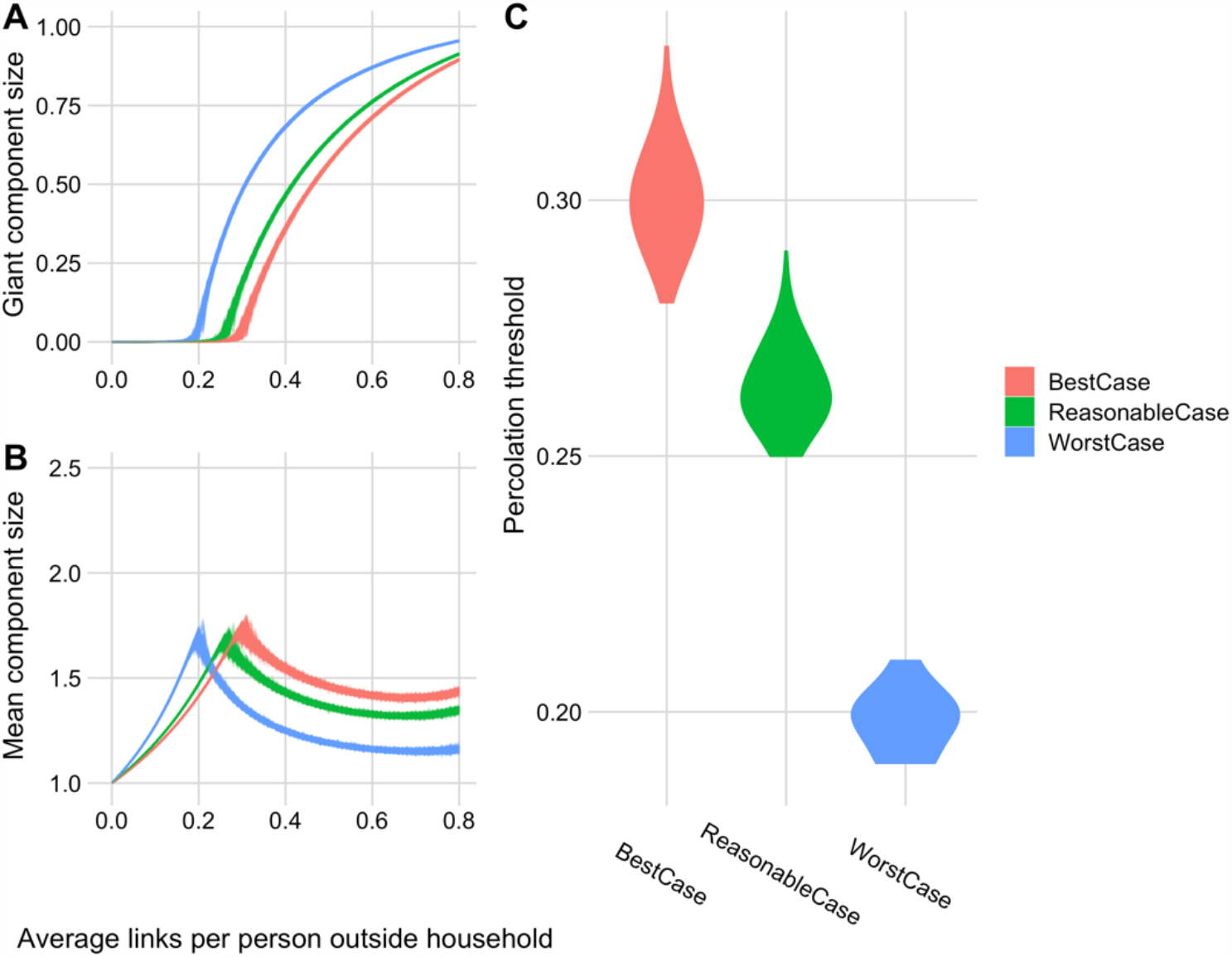
Percolation analysis for plausible bubbling scenarios. A) The proportion of households connected to the giant component. B) The mean component size (order parameter) for the same bubbling strategies. C) The percolation threshold for the different bubbling assumptions.

**Figure 4:**
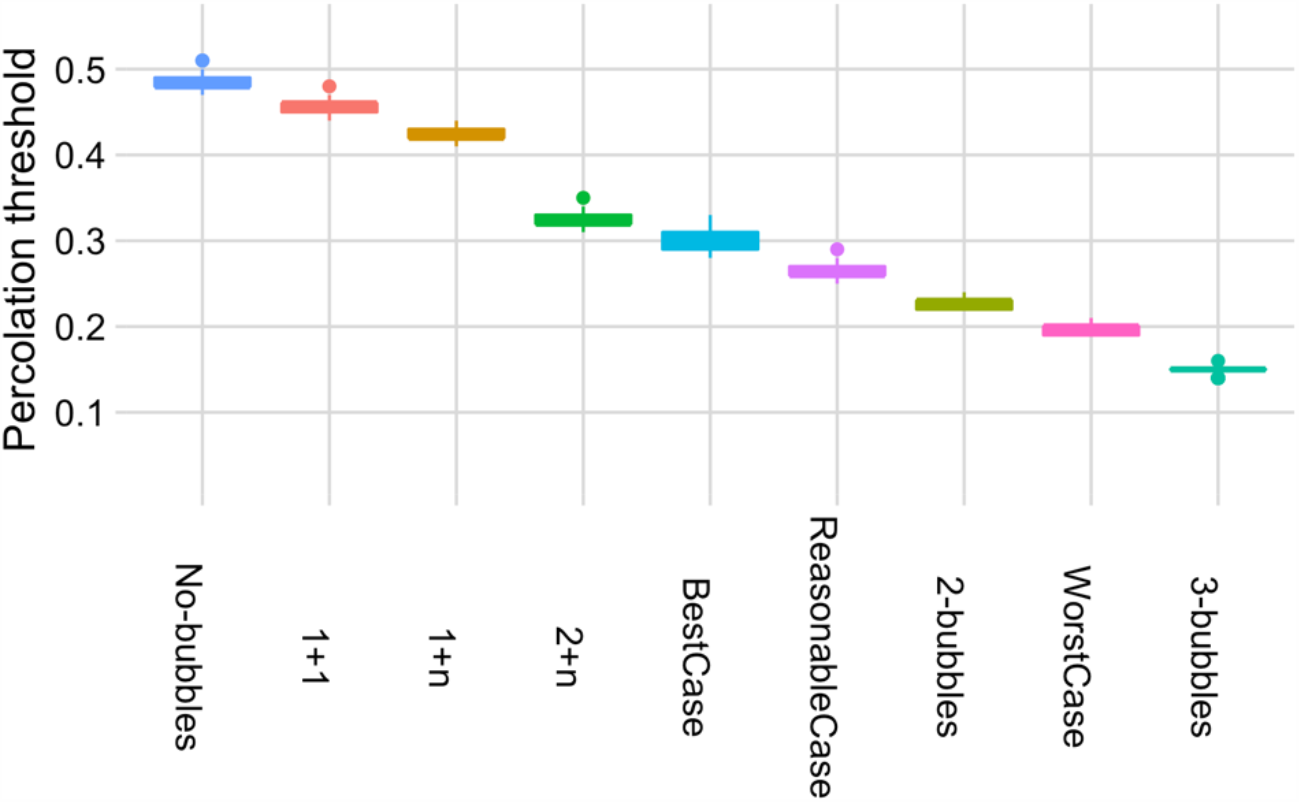
The percolation threshold for all bubbling scenarios ordered from most vulnerable to link removal (left) to least vulnerable (right).

**Figure 5:**
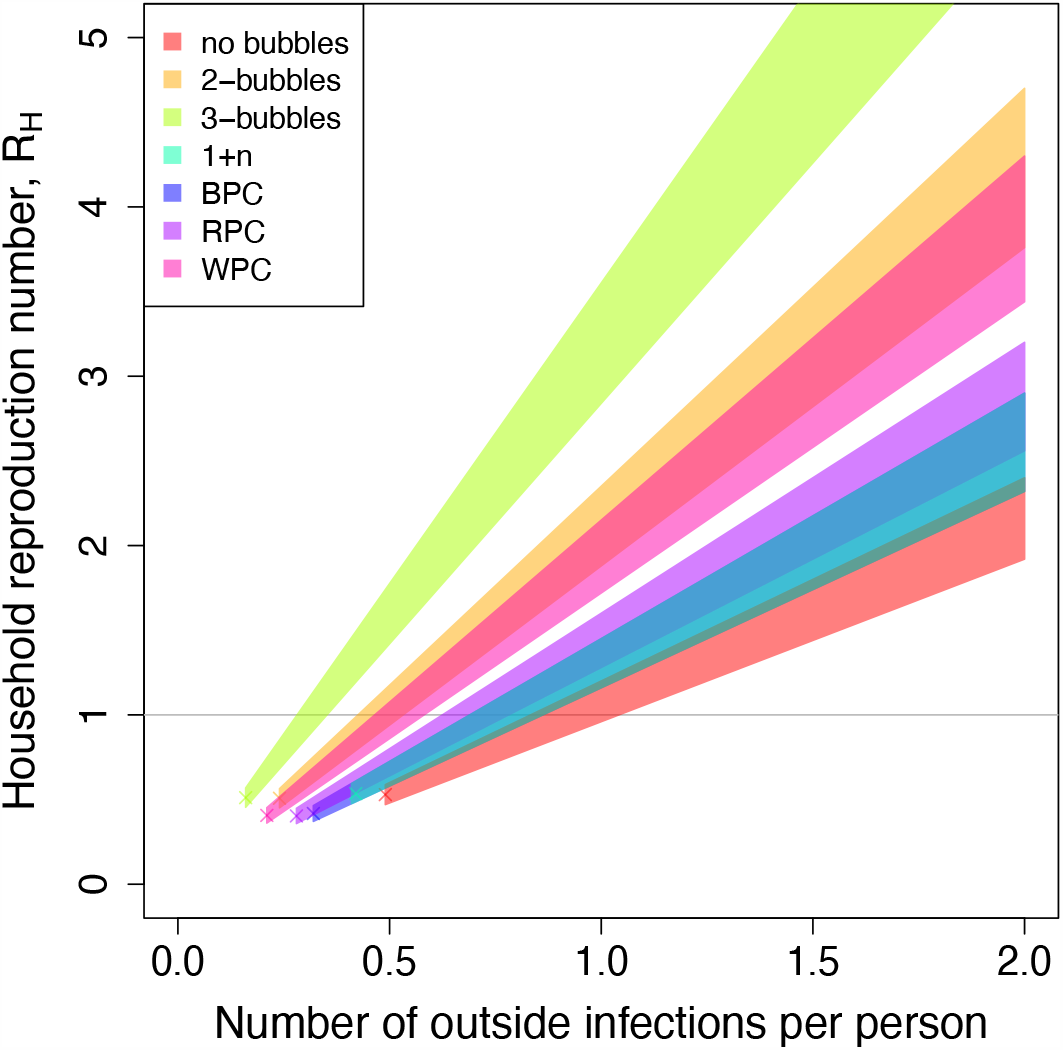
Household reproduction number for bubbling scenarios as a function of the number of outside infections per person.

Single-person households (households of size 1) joining with another single-person household (scenario 1+1), or another household of random size (scenario 1+n) has a relatively modest impact on network connectivity and transmission. The average bubble size increases by 0.4 and 0.5 persons (Figure 2). The percolation threshold is reduced by less than 15%. This translates to an increase in the household reproduction number of less than 0.3, and in practice this difference might not be observable with heterogeneity in household secondary attack rates (Figure 5 and Table 1).

**Table 1:** Comparison of bubbling scenarios

| | Average bubble size | Theoretical $p_c$ | Measured $p_c$ | $R_H$ when $p_c = 1$ |
| --- | --- | --- | --- | --- |
| **No bubbles** | 2.4 | 0.476 | 0.48 | 0.96-1.2 |
| **2-bubbles** | 4.7 | 0.224 | 0.23 | 1.88-2.35 |
| **3-bubbles** | 7.1 | 0.146 | 0.15 | 2.84-3.55 |
| **1+1** | 2.8 | 0.448 | 0.44 | 1.12-1.4 |
| **1+n** | 2.9 | 0.414 | 0.41 | 1.16-1.45 |
| **2+n** | 3.9 | 0.319 | 0.33 | 1.56-1.95 |
| **Best plausible case** | 2.9 | 0.290 | 0.30 | 1.16-1.45 |
| **Reasonable plausible case** | 3.2 | 0.255 | 0.26 | 1.28-1.6 |
| **Worst plausible case** | 4.3 | 0.193 | 0.19 | 1.72-2.15 |

In the scenario where all households form 2-bubbles there is substantial impact on network connectivity and transmission potential. Average bubble size increases to 4.7 persons per bubble. The percolation threshold is decreased to 0.23 (Figure 2), which means that one in four people with social contact outside their household is sufficient for the giant component to exist. Compared to no bubbling, 2-bubbles have the potential to increase network connectivity and increase the household reproduction number. For an average of one outside contact per person, the estimated household reproduction number is 1.9 - 2.4 (Figure 5 and Table 1).

All households forming 3-bubbles is the worst scenario we considered. As for 2-bubbles, the mean bubble size is increased to 7.1. The percolation threshold is 0.15, which means that a giant component forms if 1 in 6 persons have an external social contact. For an average of one outside contact per person, the estimated household reproduction number is 2.84 - 3.55.

For plausible scenarios with variable uptake of bubbling and heterogeneity in bubble sizes, results are considerably less dramatic than when all households form a bubble. The best plausible case, with one third of households forming 2-household and 3-household bubbles in approximately equal proportions, results in household-to-household transmission potential similar to 1+n bubbling. The reasonable plausible scenario where 50% of households for a bubble of 2 or 3 households results in network connectivity and transmission potential that is slightly greater than the best plausible case, however the difference is relatively modest and might be unobservable for heterogeneity in secondary attack rates. The impact of reasonable plausible bubbling compared to no-bubbling could be mitigated by individuals reducing their external contact rate by 0.25, for example a bubble of four individuals could reduce their external contacts from four to three.

Finally, the worst plausible scenario, where 75% of households form a bubble, produces similar characteristics to the 2-bubble scenario. Although the average bubble size is slightly smaller (4.3 for the worst plausible case versus 4.7 to 2-bubbles), the percolation threshold is lower for the worst plausible case due to the occurrence of large bubbles which link up the network (see Figure 3 and Table 1).

## Discussion

In this analysis, we provide a simple network framework in which to quantify the effect of household bubbles on the transmission of COVID-19. We find that the creation of support bubbles between a single-person household and another household of any size has a small impact on transmission. The ubiquitous generation of bubbles for all households has the potential to make transmission extremely hard to control, therefore household bubbles should not be encouraged in general. However, we find that for intermediate uptake rates - for example where 50% of households form a temporary “Christmas” bubble - that the additional transmission potential could be mitigated by a reduction in contacts outside the home.

There are natural parallels between network theory and disease dynamics, with analogies between the link probability and the giant component and the reproduction number and final epidemic size. Percolation theory has been used before to describe disease transmission in structured populations (4,9), although often using a theoretical framework, rather than to inform interventions (with the possible exception of (13)). With regards to the impact of bubbling, our results concord with Leng et al (2020) who concluded that bubbling of single-person households have a minimal impact on transmission (14).

There are a number of limitations to our analysis. First, we used a relatively simple network formulation in which the number of external contacts was proportional to household size. While this is true in general, in reality there are more complex patterns where the number of external contacts saturates with household size. Second, the model had no time dependence, therefore we were not able to capture the formation and dissolution of bubbles, and this is particularly relevant for temporary festive bubbles. Third, the model contains no spatial component. It is likely that forming local bubbles is preferable to long-distance bubbles, but we were not able to investigate that question here. Fourth, we were limited by a lack of data on current bubbling practices. The ONS Opinions and Lifestyle survey (15) reports that around 40% of adults in the UK have formed a support bubble in 2020, but we do not have data broken down by household size or on future bubbling intentions. Our estimates of the household reproduction number are dependent on the secondary attack rate in households, which is also uncertain and probably varies with household size.

This work provides quantitative insight into the impact of bubbling on the transmission dynamics of COVID-19. We find that, in a UK setting, the formation of bubbles can be detrimental if taken up by a large proportion of the population. Therefore, messaging around bubbling should be framed in a way that communicates the negative implications as well as the benefits. In particular, large gatherings of many households should be discouraged unless absolutely necessary. It is not clear how these conclusions would translate to other settings, where the definition of a household, the number of people per household and the out-of-household social mixing vary considerably. Future work could use data from other countries to explore the implications of bubbles for different social settings.

## Data Availability

All data used in the paper are publicly available through the Office for National Statistics websites.

## Acknowledgements

We thank member of SPI-M for useful suggestions and comments and Joseph Hilton for providing the household data in a convenient manner and Thomas House and Lorenzo Pellis for instructive comments and discussions. LD and EBP are supported by Medical Research Council (MRC) (MC/PC/19067) and (MR/038613/1). LL acknowledges the financial support of the EPSRC via Early Career Fellowship EP/P01660X/1.

## Footnotes

1 Indeed, in the thermodynamic limit, when the number of nodes becomes very large, the order parameter diverges(2).

